# Endocrine therapy use and the risk of cardiovascular disease in postmenopausal breast cancer survivors: two cohort studies in the UK and US

**DOI:** 10.1101/19010223

**Authors:** Anthony Matthews, Sharon Peacock Hinton, Susannah Stanway, Alexander R Lyon, Liam Smeeth, Jennifer L. Lund, Krishnan Bhaskaran

**Author notes:** Equal contribution (Jennifer L. Lund, Krishnan Bhaskaran). **Corresponding author:** Anthony Matthews: Unit of Epidemiology, Institute of Environmental Medicine, Karolinska Institutet, 171 77 Stockholm, Sweden.

## Abstract

**Objective:** Examine the effect of tamoxifen and aromatase inhibitors on 12 clinically relevant individual cardiovascular outcomes in postmenopausal female breast cancer survivors using large-scale datasets from the UK and US.

**Design:** Two prospective cohort studies

**Setting:** Population-based using data from the UK Clinical Practice Datalink linked with Hospital Episode Statistics (2002-2016), and the US Surveillance, Epidemiology and End Results-Medicare database (2008-2013).

**Participants:** 10005 and 22027 postmenopausal women with breast cancer in the UK and US respectively.

**Exposures:** Aromatase inhibitor compared with tamoxifen use; the US cohort additionally included a comparison with an “unexposed” group of women with oestrogen or progesterone receptor positive breast cancer but no endocrine therapy use.

**Outcomes:** 12 clinically relevant individual cardiovascular outcomes (and two composite coronary and venous thromboembolic outcomes)

**Results:** In both the UK and the US, there was evidence of an increased risk of coronary artery disease in aromatase inhibitor compared with tamoxifen users (UK incidence rate: 10.18 vs 6.87 per 1000 person-years, HR: 1.29, 0.94-1.76; US incidence rate: 35.26 vs 26.95 per 1000 person-years, HR: 1.29, 1.06-1.55), but the US data showed no increase in risk compared with the unexposed group (incidence rate for tamoxifen vs unexposed: 26.95 vs 38.70 per 1000 person-years, HR: 0.74, 0.60-0.92; incidence rate for aromatase inhibitors vs unexposed: 35.26 vs 28.70, HR: 0.96, 0.83-1.10). Similar patterns were seen for other cardiovascular outcomes such as arrhythmia, heart failure, and valvular heart disease. As expected, there were more venous thromboembolic events in tamoxifen users compared with both aromatase inhibitor users and those unexposed. There was a high degree of consistency between results in the two countries.

**Conclusions:** Increased risks of several cardiovascular diseases among aromatase inhibitor compared with tamoxifen users appeared to be driven by protective effects of tamoxifen, rather than toxic effects of aromatase inhibitors. We also confirmed the known increased risk of venous thromboembolic events in tamoxifen users.

**WHAT THIS PAPER ADDS:** *What is already known on this topic:*

- It is known that tamoxifen use increases venous thromboembolism risk, but evidence for other cardiovascular outcomes is less clear.
- Patterns of results are suggestive of a lower risk of coronary heart disease outcomes with tamoxifen compared to both aromatase inhibitor use and no tamoxifen or placebo, but cardiovascular events are often a secondary consideration and inconsistently reported in trials, and most observational studies use composite cardiovascular definitions, ignoring potentially differential effects on specific cardiovascular outcomes.

*What this study adds:*

- Among postmenopausal women with breast cancer, we found an increased risk of several cardiovascular diseases in aromatase inhibitor compared with tamoxifen users across two countries, which appeared to be driven by protective effects of tamoxifen, rather than toxic effects of aromatase inhibitors. We also found the known increased venous thromboembolism risk in tamoxifen users.
- There was no evidence that aromatase inhibitors or tamoxifen increases cardiovascular disease risk, other than the known increased venous thromboembolism risk with tamoxifen use. However, there was an apparent protective effect of tamoxifen on other cardiovascular outcomes.

## BACKGROUND

Breast cancer is the most common cancer among women worldwide; 83% of breast cancer patients are diagnosed with oestrogen or progesterone receptor positive disease (ER/PR+) and are recommended endocrine therapies to minimise risk of recurrence.[1] Tamoxifen reduces the rate of breast cancer recurrence by nearly half, regardless of menopausal status,[2] but since 2006 aromatase inhibitors (AI) are also recommended to postmenopausal women due to greater efficacy over tamoxifen.[3] However, it is hypothesised that these drugs may affect cardiovascular disease (CVD) risk through pathways including suppression of protective effects of circulating oestrogen,[4] changes in cholesterol,[5, 6] and reduction of antithrombin levels.[7]

A recent systematic review collated all evidence for the risk of CVD for users of tamoxifen, AIs, and the comparative risk between the two drugs.[8] It suggested that tamoxifen use increases risk of venous thromboembolic events, but evidence for other outcomes was less clear. Patterns of results are suggestive of a lower risk of coronary heart disease outcomes with tamoxifen compared to both AI use and no tamoxifen or placebo, but CVD events are often a secondary consideration and inconsistently reported in trials, and most observational studies use composite CVD definitions, ignoring potentially differential effects on specific CVDs.

Given the uncertainty and clinical importance, we aimed to examine the effects of tamoxifen and AIs on 12 clinically relevant individual CVD outcomes (and two composite outcomes) in postmenopausal female breast cancer survivors using two large-scale datasets from both the UK and US.

### METHODS

### Study design and data sources

We assembled two separate and nationally representative cohorts of women with incident postmenopausal breast cancer using prospectively collected data from large UK and US electronic healthcare databases. In the UK, we used Clinical Practice Research Datalink primary care data (CPRD GOLD) and linked Hospital Episode Statistics (HES). [9] In the US, we used the Surveillance, Epidemiology, and End Results program (SEER) and Medicare linked database.[10] Full detail on these data sources are available in Appendix 1.

### Study populations

Differences between the study populations were largely driven by differences in data available within the two *cohorts*, which are fully explained in Appendix 2.

#### UK cohort

We identified women with linked CPRD and HES data over 54 years (median age of the menopause in Europe [11]), with incident breast cancer in CPRD (after at least one year of CPRD follow-up), who initiated AIs or tamoxifen in primary care after their diagnosis, from 1^st^ January 2002 (date from which preliminary analysis showed that third generation AIs came into widespread use) to 31^st^ March 2016 (latest CPRD and HES linkage). Follow-up began at the latest of one year after breast cancer diagnosis, or first AI or tamoxifen prescription (hereafter index date). Women were excluded if prior to their index date they: died, transferred out of the CPRD, had any other cancer diagnosis, or were diagnosed with the CVD event of interest (at any point prior to index date).

#### US cohort

We identified women over 65 years with incident ER/PR+ and stage 1-3 breast cancer and continuous Medicare Parts A, B and D enrolment (and no managed care coverage) for the 12-months prior to the month of cancer diagnosis from 1^st^ January 2008 (Medicare Part D data are available from 2007) and 31^st^ December 2013 (last capture of cancer cases in SEER). Women with an endocrine therapy prescription prior to their breast cancer diagnosis were excluded. Follow up began one year after the date of breast cancer (hereafter index date). Women were excluded if prior to their index date they: died, discontinued from Medicare Parts A, B, or D, had any other cancer diagnosis, or were diagnosed with the CVD event of interest (within a 3-year look back period to ensure likelihood of capturing a prior event was not dependent on age as Medicare follow up starts at 65 years).

### Exposure, outcomes, and covariates

Tamoxifen and AI exposures were identified using prescription codes in the UK (available at https://doi.org/10.17037/DATA.177), and National Drug Codes (NDCs) and Healthcare Common Procedural Coding System (HCPCS) procedure codes in the US (Appendix 3). The primary exposure was ever use of tamoxifen, ever use of AI, or ever use of both drugs; the US study included an additional category of no exposure to any endocrine therapy (which did not exist in the UK study, because receipt of endocrine therapy was an inclusion criterion). Exposure was time-updated to indicate a woman had been exposed to both drugs if they switched endocrine therapies during follow-up. The baseline exposure group was classified as ever exposure to tamoxifen for the ever AI vs tamoxifen analyses in both the UK and US, whereas the baseline exposure group was changed to no exposure to any endocrine therapy for the ever AI/tamoxifen vs unexposed analyses that were only possible in the US. In secondary analyses, we considered time-updated current exposure to endocrine therapy, categorised as current tamoxifen use, current AI use, no current therapy and prior AI use, no current therapy and prior tamoxifen use only, and (in the US study only) no current or past endocrine therapy. A drug exposure was continuous if a further prescription for the same endocrine therapy followed within 30 days of the end of the previous prescription date plus the days of drug supplied. Further information is in Appendix 4, and visualisations of exposure categorisations in Appendices 5 and 6.

The main CVD outcomes were: coronary artery disease (angina, myocardial infarction (MI), revascularisation procedures, sudden cardiac arrest (SCA)); peripheral vascular disease (PVD); stroke; arrhythmia; heart failure (HF, including cardiomyopathy); pericarditis, valvular heart disease (VHD); and venous thromboembolism (VTE) (deep vein thrombosis (DVT), pulmonary embolism (PE)). Composite CVD outcomes and individual components of the composite outcomes were analysed separately. Events were identified through clinical diagnoses using NHS Read codes in the CPRD and International Classification of Disease (ICD), 10^th^ edition codes in HES in the UK (available at https://doi.org/10.17037/DATA.177), and ICD-9 and Healthcare Common Procedure Coding System (HCPCS) codes in the US study (Appendix 7).

In both studies, we adjusted for age, cardiovascular history and risk factors, use of cardio-protective medications, other comorbidities, time since index date, and calendar year. In the UK we were additionally able to adjust for smoking, alcohol use, body mass index, and index of multiple deprivation (a measure of socioeconomic status); in the US study we were additionally able to adjust for race, region and use of anti-cancer therapies. A full comparison of the covariates considered in the UK and US is in Appendix 8. Algorithms to define confounders in the UK are in Appendix 9, and code lists used for variable definitions are at https://doi.org/10.17037/DATA.177. Codes used to identify prescriptions in the US are in Appendix 10.

### Statistical Analysis

Observation time began at index date and ended at the earliest of: a CVD event of interest; diagnosis of another (non-breast) cancer; death; transfer out of CPRD/end of Medicare Parts A, B, or D enrolment; or end of study period (31^st^ March 2016 in UK which is the last available CPRD collection date, and 31^st^ December 2014 in US which is the last available Medicare collection date).

Separate analyses were conducted in UK and US data. Distributions of baseline characteristics of patients at index date were described. Number of events and crude incident rates of each outcome of interest by exposure were calculated. Cox proportional hazards regression models with an underlying age timescale were fitted for each outcome of interest to obtain unadjusted hazard ratios (HRs) and 95% CIs for the association between endocrine therapy use and outcome; those with a diagnosis of the specific outcome of interest before the index date were excluded from the analysis for that outcome. All covariates were then added to obtain fully-adjusted HRs and 95% CIs. The analysis was repeated for the primary and secondary exposure definitions. Using the primary exposure definitions in each country, we then fitted interactions to investigate effect modification by current age (54-69, 70+ in the UK; 66-84, 85+ in the US); time since index date (0-1 years, 1-3 years, 3+ years); and history of any CVD prior to index date for the coronary artery disease (composite), arrhythmia, stroke, pericarditis (US only), heart failure, VHD, and VTE (composite) outcomes (interactions were not investigated for other outcomes due to limited power). Women with missing data (8.7% in the UK, and 5.1% in the US) were excluded from all analyses (complete case analysis), which is valid in a regression context if missingness is conditionally independent of the outcome.[12]

### Sensitivity analyses

To ensure comparability between the UK and US, the main analyses were repeated with the study populations and covariates modified to be as similar as possible. Further details in Appendix 11. Furthermore, in case of misclassification of exposure, the grace period used to define a continuous prescription was extended from 30 days to 3 months, 6 months, and 1 year in the analyses of current endocrine therapy exposure. Finally, a post-hoc quantitative bias analysis explored potential unmeasured confounding in the large estimated protective effect of tamoxifen use on the risk of MI (Appendix 12).[13]

Patients and the public were not involved at any point during this research.

## RESULTS

The UK study included 10005 women, with 4716 (47%) initially prescribed tamoxifen and 5289 (53%) initially prescribed an AI (Table 1, flow diagram in Figure 1). Mean follow-up from index date for individual CVD outcomes ranged from 4-4.1 years. The US study included 22027 women, with 4667 (22%), 2286 (10%) and 15074 (68%) initially filling no endocrine therapy, tamoxifen, and an AI respectively (Table 2, flow diagram in Figure 1). Mean follow-up from index date for individual CVD outcomes ranged from 2.3-2.5 years.

**Table 1.** Characteristics of study population based on their initial exposure in the UK

|  | **Tamoxifen** | **AI** | **Total** |
| --- | --- | --- | --- |
| **N** | 4716 (100) | 5289 (100) | 10005 (100) |
| **Age (yrs)** |  |  |  |
| 54-59 | 911 (19.3) | 716 (13.5) | 1627 (16.3) |
| 60-69 | 1850 (39.2) | 1972 (37.3) | 3822 (38.2) |
| 70+ | 1955 (41.5) | 2601 (49.2) | 4556 (45.5) |
| Median (IQR) | 68 (62-76) | 70 (63-79) | 69 (62-78) |
| **Year of breast cancer** |  |  |  |
| 2002 | 579 (12.3) | 72 (1.4) | 651 (6.5) |
| 2003 | 605 (12.8) | 123 (2.3) | 728 (7.3) |
| 2004 | 571 (12.1) | 186 (3.5) | 757 (7.6) |
| 2005 | 512 (10.9) | 289 (5.5) | 801 (8) |
| 2006 | 467 (9.9) | 370 (7) | 837 (8.4) |
| 2007 | 386 (8.2) | 448 (8.5) | 834 (8.3) |
| 2008 | 380 (8.1) | 495 (9.4) | 875 (8.7) |
| 2009 | 290 (6.1) | 533 (10.1) | 823 (8.2) |
| 2010 | 223 (4.7) | 586 (11.1) | 809 (8.1) |
| 2011 | 216 (4.6) | 619 (11.7) | 835 (8.3) |
| 2012 | 210 (4.5) | 539 (10.2) | 749 (7.5) |
| 2013 | 174 (3.7) | 504 (9.5) | 678 (6.8) |
| 2014 | 93 (2) | 451 (8.5) | 544 (5.4) |
| 2015 | 10 (.2) | 74 (1.4) | 84 (.8) |
| **BMI (kg/m2)** |  |  |  |
| <18 | 59 (1.3) | 63 (1.2) | 122 (1.2) |
| 18-24 | 1693 (35.9) | 1619 (30.6) | 3312 (33.1) |
| 25-29 | 1549 (32.8) | 1801 (34.1) | 3350 (33.5) |
| 30-34 | 800 (17) | 979 (18.5) | 1779 (17.8) |
| ≥35 | 345 (7.3) | 548 (10.4) | 893 (8.9) |
| Missing | 270 (5.7) | 279 (5.3) | 549 (5.5) |
| Median (IQR) | 26 (23-30) | 27 (24-31) | 27 (24-31) |
| **Smoking status** |  |  |  |
| Never smoker | 2423 (51.4) | 2517 (47.6) | 4940 (49.4) |
| Current smoker | 503 (10.7) | 482 (9.1) | 985 (9.8) |
| Ex-smoker | 1761 (37.3) | 2268 (42.9) | 4029 (40.3) |
| Missing | 29 (.6) | 22 (.4) | 51 (.5) |
| **Alcohol use** |  |  |  |
| Non drinker | 618 (13.1) | 613 (11.6) | 1231 (12.3) |
| Current | 3320 (70.4) | 3628 (68.6) | 6948 (69.4) |
| Ex-drinker | 480 (10.2) | 715 (13.5) | 1195 (11.9) |
| Missing | 298 (6.3) | 333 (6.3) | 631 (6.3) |
| **Systolic BP** |  |  |  |
| Low/ideal | 530 (11.2) | 599 (11.3) | 1129 (11.3) |
| Pre-high | 1862 (39.5) | 2327 (44) | 4189 (41.9) |
| High | 2314 (49.1) | 2355 (44.5) | 4669 (46.7) |
| Missing | 10 (.2) | 8 (.2) | 18 (.2) |
| **Diastolic BP** |  |  |  |
| Low/ideal | 2130 (45.2) | 2650 (50.1) | 4780 (47.8) |
| Pre-high | 1988 (42.2) | 2058 (38.9) | 4046 (40.4) |
| High | 588 (12.5) | 573 (10.8) | 1161 (11.6) |
| Missing | 10 (.2) | 8 (.2) | 18 (.2) |
| **IMD category** |  |  |  |
| 1 | 870 (18.4) | 962 (18.2) | 1832 (18.3) |
| 2 | 943 (20) | 1214 (23) | 2157 (21.6) |
| 3 | 925 (19.6) | 1054 (19.9) | 1979 (19.8) |
| 4 | 1052 (22.3) | 936 (17.7) | 1988 (19.9) |
| 5 | 926 (19.6) | 1122 (21.2) | 2048 (20.5) |
| Missing | 0 (0) | 1 (0) | 1 (0) |
| **CVD related treatment before index** |  |  |  |
| Statins | 1100 (23.3) | 1903 (36) | 3003 (30) |
| ACEI | 1195 (25.3) | 1823 (34.5) | 3018 (30.2) |
| CCB | 1195 (25.3) | 1764 (33.4) | 2959 (29.6) |
| ARB | 493 (10.5) | 795 (15) | 1288 (12.9) |
| Anti-platelets | 1132 (24) | 1639 (31) | 2771 (27.7) |
| **Comorbidities before index** |  |  |  |
| RA | 138 (2.9) | 137 (2.6) | 275 (2.7) |
| Diabetes | 462 (9.8) | 728 (13.8) | 1190 (11.9) |
| CKD | 867 (18.4) | 1090 (20.6) | 1957 (19.6) |
| **CVD before index** |  |  |  |
| Non-venous CVD | 1031 (21.9) | 1636 (30.9) | 2667 (26.7) |
| VTE before index | 144 (3.1) | 324 (6.1) | 468 (4.7) |

**Table 2.** Characteristics of study population based on their initial exposure in the US

|  | Unexposed | Tamoxifen | AI | Total |
| --- | --- | --- | --- | --- |
| N | 4667 (100) | 2286 (100) | 15074 (100) | 22027 (100) |
| **Age at index date (yrs)** |  |  |  |  |
| 66-74 | 1538 (33) | 897 (39.2) | 7505 (49.8) | 9940 (45.1) |
| 75-84 | 1937 (41.5) | 994 (43.5) | 5894 (39.1) | 8825 (40.1) |
| 85+ | 1192 (25.5) | 395 (17.3) | 1675 (11.1) | 3262 (14.8) |
| Median (IQR) | 79 (73-85) | 77 (72-83) | 75 (71-81) | 76 (71-82) |
| **Ethnicity** |  |  |  |  |
| White | 4002 (85.8) | 2028 (88.7) | 12782 (84.8) | 18812 (85.4) |
| Black | 360 (7.7) | 102 (4.5) | 1099 (7.3) | 1561 (7.1) |
| Other | 93 (2) | 47 (2.1) | 335 (2.2) | 475 (2.2) |
| Asian | 123 (2.6) | 68 (3) | 498 (3.3) | 689 (3.1) |
| Hispanic | - | - | - | 397 (1.8) |
| Native American | - | - | - | 52 (.2) |
| Missing | - | - | - | 41 (.2) |
| **SEER Region** |  |  |  |  |
| North East | 760 (16.3) | 283 (12.4) | 3320 (22) | 4363 (19.8) |
| South | 1023 (21.9) | 605 (26.5) | 3778 (25.1) | 5406 (24.5) |
| North Central | 695 (14.9) | 448 (19.6) | 1761 (11.7) | 2904 (13.2) |
| West | 2157 (46.2) | 939 (41.1) | 6130 (40.7) | 9226 (41.9) |
| Missing | 32 (.7) | 11 (.5) | 85 (.6) | 128 (.6) |
| **Year of breast cancer** |  |  |  |  |
| 2008 | 847 (18.1) | 414 (18.1) | 2075 (13.8) | 3336 (15.1) |
| 2009 | 831 (17.8) | 449 (19.6) | 2140 (14.2) | 3420 (15.5) |
| 2010 | 729 (15.6) | 368 (16.1) | 2394 (15.9) | 3491 (15.8) |
| 2011 | 744 (15.9) | 333 (14.6) | 2625 (17.4) | 3702 (16.8) |
| 2012 | 756 (16.2) | 350 (15.3) | 2709 (18) | 3815 (17.3) |
| 2013 | 760 (16.3) | 372 (16.3) | 3131 (20.8) | 4263 (19.4) |
| **Stage of breast cancer** |  |  |  |  |
| Stage I | 3034 (65) | 1486 (65) | 8379 (55.6) | 12899 (58.6) |
| Stage II | 1275 (27.3) | 660 (28.9) | 5267 (34.9) | 7202 (32.7) |
| Stage III | 358 (7.7) | 140 (6.1) | 1428 (9.5) | 1926 (8.7) |
| **Grade of breast cancer** |  |  |  |  |
| 1 | 1522 (32.6) | 765 (33.5) | 4273 (28.3) | 6560 (29.8) |
| 2 | 2071 (44.4) | 1109 (48.5) | 7350 (48.8) | 10530 (47.8) |
| 3 | 853 (18.3) | 324 (14.2) | 2810 (18.6) | 3987 (18.1) |
| Missing | 221 (4.7) | 88 (3.8) | 641 (4.3) | 950 (4.3) |
| **Cancer treatments** |  |  |  |  |
| Taxane | 570 (12.2) | 162 (7.1) | 2415 (16) | 3147 (14.3) |
| Anthracyclines | 259 (5.5) | 68 (3) | 820 (5.4) | 1147 (5.2) |
| Trastuzumab | 226 (4.8) | 39 (1.7) | 687 (4.6) | 952 (4.3) |
| Other treatment | 753 (16.1) | 244 (10.7) | 2992 (19.8) | 3989 (18.1) |
| **Comorbidities** |  |  |  |  |
| RA | 185 (4) | 103 (4.5) | 547 (3.6) | 835 (3.8) |
| CKD | 383 (8.2) | 155 (6.8) | 1113 (7.4) | 1651 (7.5) |
| Hypertension | 3426 (73.4) | 1612 (70.5) | 11113 (73.7) | 16151 (73.3) |
| Diabetes | 1313 (28.1) | 598 (26.2) | 4545 (30.2) | 6456 (29.3) |
| **CVD related treatment before index** |  |  |  |  |
| Statins | 1778 (38.1) | 948 (41.5) | 6988 (46.4) | 9714 (44.1) |
| Hypertensives | 169 (3.6) | 83 (3.6) | 577 (3.8) | 829 (3.8) |
| ACEi | 962 (20.6) | 477 (20.9) | 3251 (21.6) | 4690 (21.3) |
| CCB | 850 (18.2) | 364 (15.9) | 2696 (17.9) | 3910 (17.8) |
| ARB | 593 (12.7) | 255 (11.2) | 2063 (13.7) | 2911 (13.2) |
| **Past CVD** |  |  |  |  |
| Non venous CVD | 2989 (64) | 1281 (56) | 8896 (59) | 13166 (59.8) |
| VTE | 162 (3.5) | 31 (1.4) | 385 (2.6) | 578 (2.6) |

\*Cell numbers within ethnicity suppressed due to some cells containing numbers $\leq 11$

**Figure 1.**
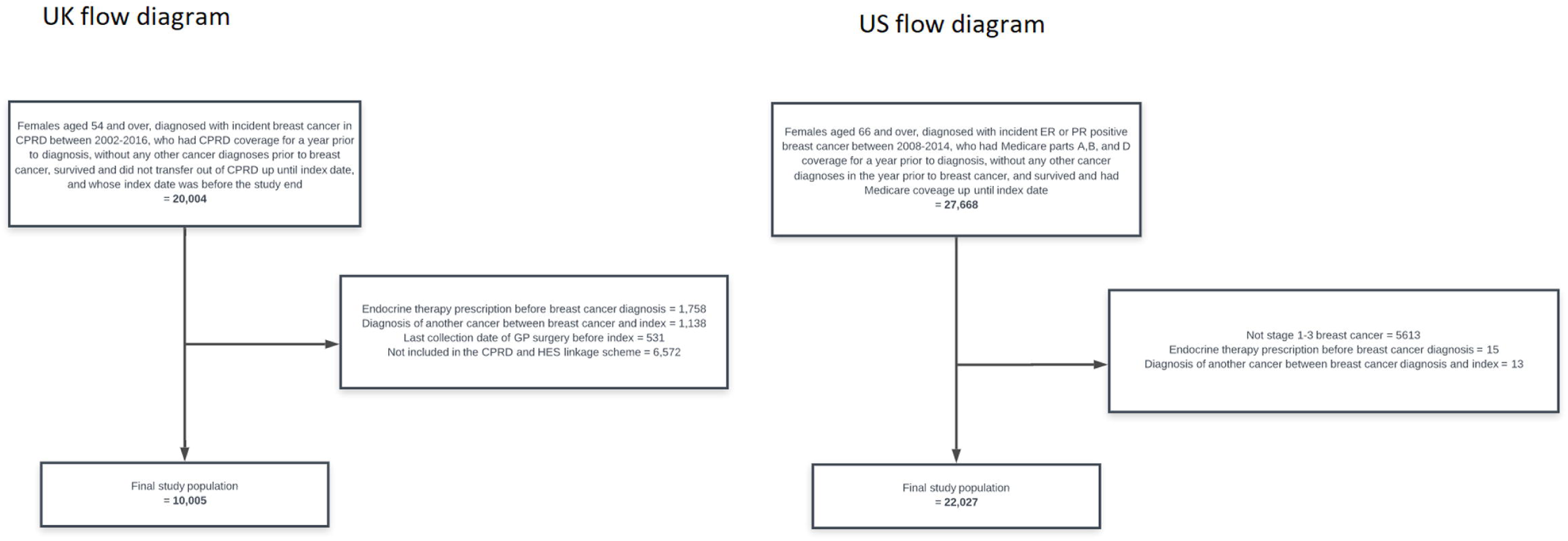
Flow diagrams of study populations in the UK and US.

### Ever Exposure analyses

#### Ever AI vs Tamoxifen use

In both the UK and US settings, there was a higher observed rate of most CVDs, excluding VTE outcomes, among those ever exposed to an AI compared with tamoxifen (Appendices 13-14). In adjusted analyses, there was strong evidence of a higher risk of heart failure in AI compared with tamoxifen users in the UK (HR: 1.68, 1.24-2.26, Figure 2), which was not replicated in the US (HR: 0.96, 0.83-1.12). Other adjusted HRs were consistent between the two settings; there was evidence in one or both settings that AI users compared with tamoxifen users had higher risk of coronary artery disease, myocardial infarction, arrhythmia, pericarditis, and VHD (HRs ranged from 1.29-3.25 in the UK, and 1.21-1.81 in the US, Figure 2) and lower risk of deep vein thrombosis (UK HR: 0.63, 0.43-0.93; US HR: 0.81, 0.50-1.09).

**Figure 2.**
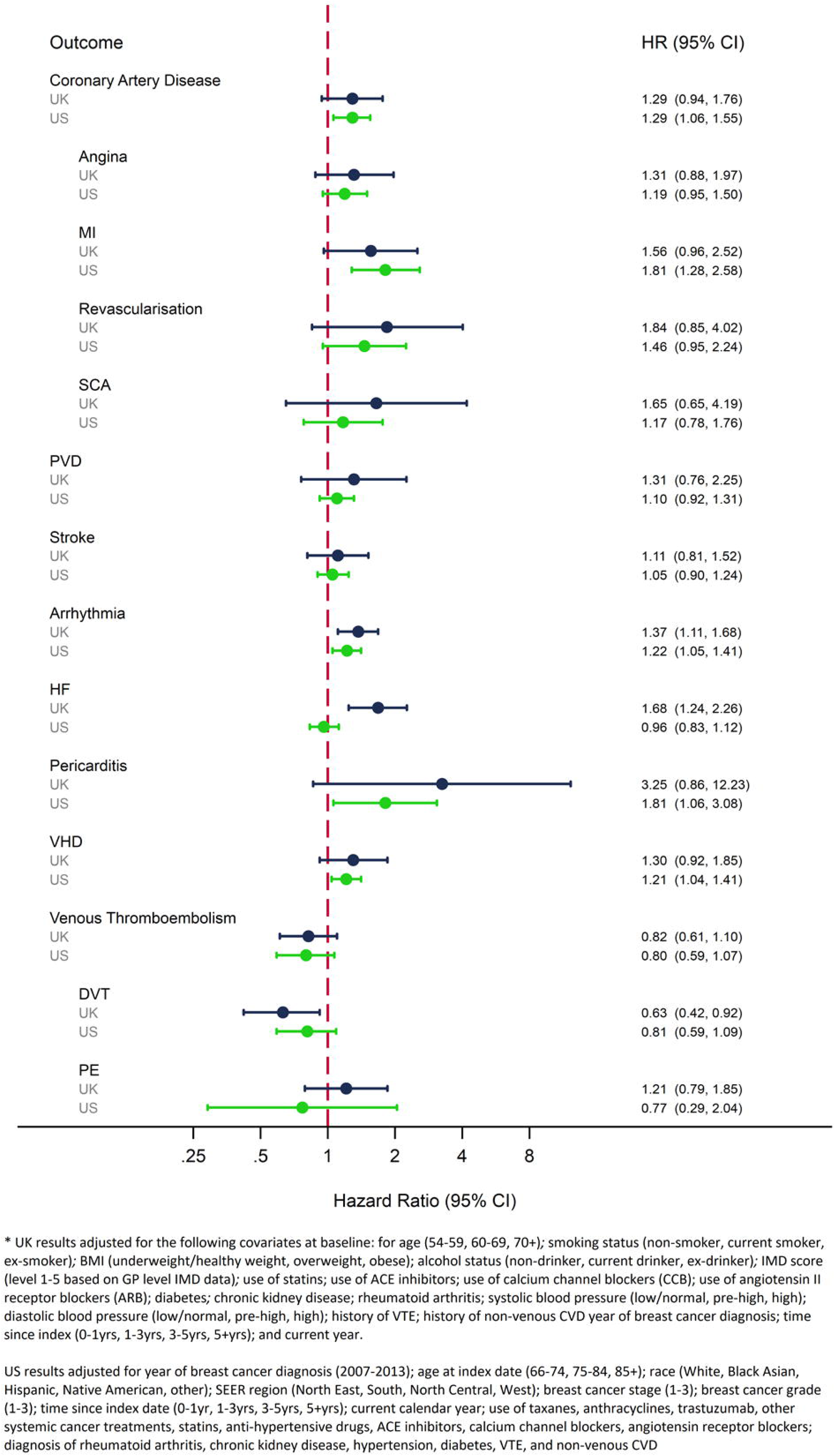
Adjusted HRs for the association between ever AI use compared with ever tamoxifen use and the risk of a range of clinical CVD outcomes in the UK and US

#### Ever AI/Tamoxifen use vs unexposed

The US study included women without exposure to either endocrine therapy; observed rates of most CVDs, excluding VTE, were lower in both the tamoxifen and AI groups compared with the unexposed group, while VTE outcome rates were higher in the endocrine therapy groups (Appendix 14). The patterns were similar in adjusted analyses (Figure 3); there was evidence that tamoxifen users had lower risks than unexposed women for coronary artery disease, myocardial infarction, stroke, arrhythmia, pericarditis, and VHD (HRs ranged from 0.37-0.87); HR point estimates for AI users vs unexposed were also in the protective direction for all non-VTE CVD outcomes, but in most cases were closer to the null than for tamoxifen, and with confidence intervals including no association. There was weak evidence of higher risk VTE in tamoxifen users compared with the unexposed (HR: 1.39, 0.98-1.98), but little evidence of a difference for AI users vs unexposed (HR: 1.14, 0.86-1.52).

**Figure 3.**
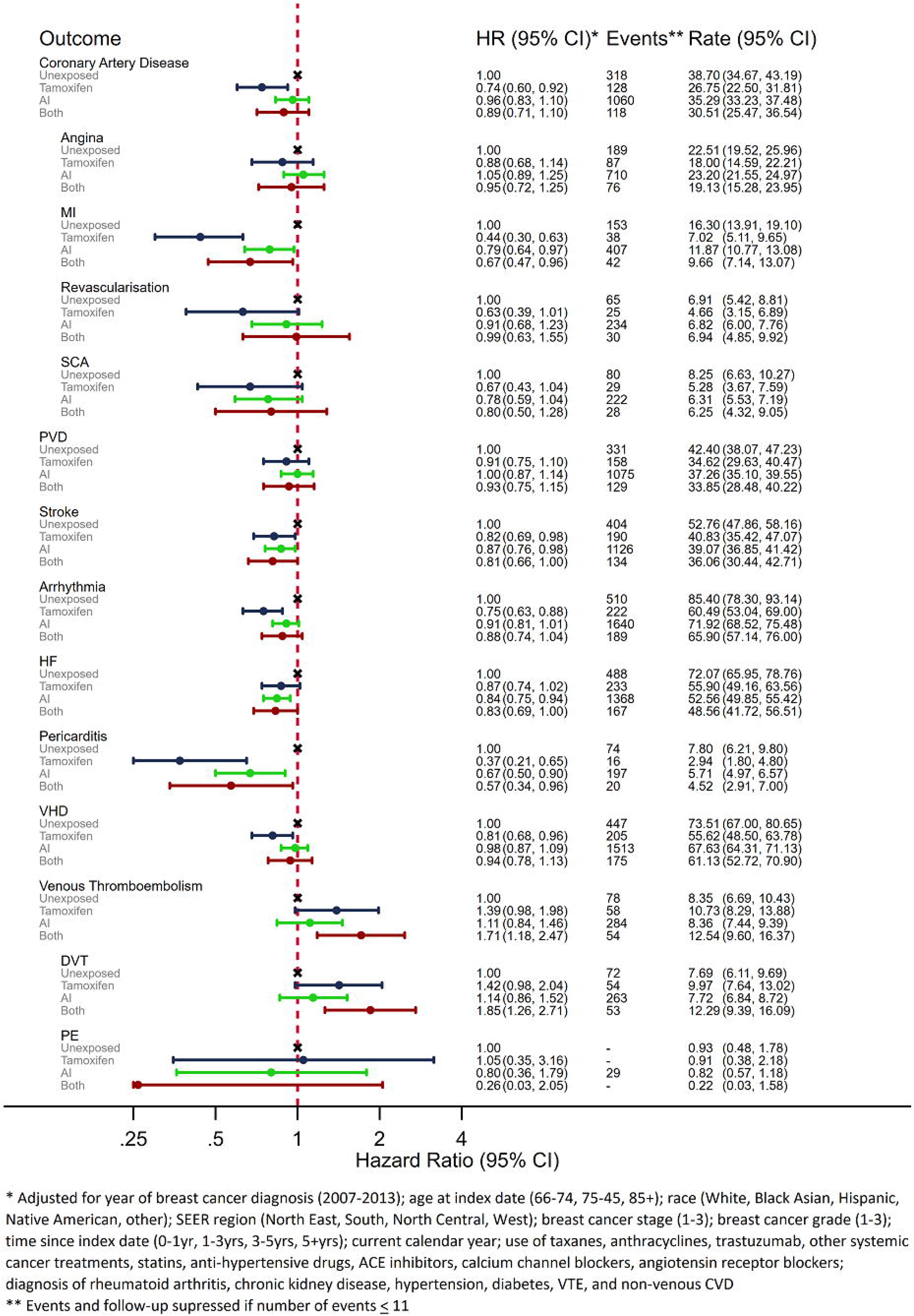
Adjusted HRs, events, and crude rate per 1000 person-years for the association between ever exposure to endocrine therapy and a range of clinical CVD outcomes in the US

### Current exposure analyses

Results for the analysis of current endocrine therapy use were consistent with the findings in the primary ever exposure analyses, and were consistent between the UK and US (Appendices 15-18).

### Effect modification

There was no strong evidence of effect modification by age, time since index date, or prior CVD in the UK or US (Appendices 19-20), though there were few events within strata, limiting precision. There was a suggestion in UK data that the raised risk of coronary artery disease in AI compared with tamoxifen users diminished over time (p=0.02), but no corresponding evidence in US data.

### Sensitivity analyses

Following modification of study populations, methodology, and covariates in the UK and US to make them as similar as possible, the risk of all CVDs associated with ever and current AI compared to tamoxifen use were generally in the same direction, albeit with less precision in UK data (Appendices 21-22). However, the discrepant results for heart failure between the two cohorts persisted.

Altering the grace period used to define continuous drug use had little effect on results (Appendices 23-24).

Quantitative bias analyses suggest that unmeasured confounding was unlikely to fully explain the large HR for the effect of ever tamoxifen use on the risk of MI (Appendix 25).

## DISCUSSION

These two population-based cohort studies using UK and US data are the first to apply similar methodology to two large populations to assess the effects of endocrine therapies on a range of CVD outcomes in postmenopausal women with breast cancer. Both countries’ results suggested a higher risk of several CVD outcomes in AI compared with tamoxifen users, with no evidence that any other characteristics, including previous CVD, modified the estimated effect. However, comparisons with women exposed to neither drug suggest that this is driven by a decreased risk of these CVD outcomes in tamoxifen users; and there was no evidence for any cardiotoxicities associated with AI use compared with those unexposed to endocrine therapy. In both the UK and the US, tamoxifen users were consistently at higher risk of VTE outcomes than both AI users and those unexposed to endocrine therapy.

The protective effect of tamoxifen on some CVDs might be explained by the drug’s effect on lipid levels. Previous studies have found tamoxifen to reduce total serum cholesterol (10-15%) and low-density lipoprotein cholesterol (15-22%).[5, 6] It has also been suggested that AIs could increase the risk of CVD compared with tamoxifen through depleting oestrogen-mediated protective CVD effects,[4] but several trials have compared hypercholesterolemia between AI and tamoxifen users, with inconclusive results.[14-16]

### Comparison to other studies

A recent meta-analysis of trials reported an increased risk of a composite CVD outcome, excluding VTE, in tamoxifen compared with AI users (RR: 1.18, 1.05-1.33),[17] with results suggestive of a cardio-protective effect of tamoxifen, consistent with the results of this study.

Several studies have focused on more specific CVD outcomes, and MI, stroke, heart failure, and VTE are the outcomes with the largest body of previous evidence, but there is little evidence for the other specific CVD outcomes that we explored. Most (4/5) previous studies directly comparing the risk of MI in AI and tamoxifen users have reported an increased risk in AI users, similar to our results (RRs ranged from 0.99-2.02).[18-22] Two previous trials and five observational studies have compared MI risk in tamoxifen users with non-use/placebo, with 4/7 studies finding a reduced risk in tamoxifen users (RRs ranged from 0.20-0.83);[23-29] two analogous studies of AI use versus placebo or non-use found no association.[16, 28] Six studies (five trials and one observational) directly compared the risk of stroke in AI and tamoxifen users, but associations in both directions have been reported.[20, 22, 30-33] Three out of five studies (one trial and four observational) comparing tamoxifen use with non-use/placebo found a protective association with stroke, as in our study (RRs ranged from 0.52-0.81).[23, 26-28, 34] In the present analysis, we found AI users to be at reduced risk of stroke compared to unexposed women; one previous study found a similar association but a second reported the opposite.[16, 28]. Two previous studies (one trial and one observational) have reported results for the comparison between AI and tamoxifen use on the risk of heart failure, [30, 33] for which we reported discrepant results between the UK and US. One reported an increased risk in AI users and the other reported no association. The discrepant results in our study could be due to residual confounding by variables not available in both datasets (cancer therapies in the UK, and lifestyle factors in the US), or the nature of the data sources (routinely collected records in the UK and claims in the US).

Six previous trials compared the risk of VTE in AI and tamoxifen users, with five reporting a decreased risk in AI compared with tamoxifen users (RR ranged from 1.25-0.61), as reported in both UK and US data in this study.[18, 20, 22, 30, 35, 36] Six out of eight studies (five trials and three observational) also reported an effect estimate that suggested an increased risk in tamoxifen users (RRs ranged from 1.64-7.10), which is in the same direction, but larger than the effect in US data here. One observational study reported an increased risk of VTE in AI users (RR: 1.84, 1.11-3.04), but US data suggested no association.

### Strengths and limitations

A major strength of this study was the use of two large data sources from different countries, with complementary strengths and different limitations. By including data from study populations with different characteristics, we could look for consistency in observed associations, and conduct different comparisons, notably between classes, and with unexposed patients. We were also able to assess the relationships between endocrine therapy treatments and a wide range of CVD outcomes, rather than the composite and individual CVD outcomes in many previous studies. We were also able to account for several important confounders such as potentially cardio-toxic treatments (anthracyclines and trastuzumab, in US data) and lifestyle factors (BMI, smoking, and alcohol, in UK data); although no confounders materially changed the crude effect estimate when adjusted for individually in the ever exposure analyses in either the UK or US (Appendices 26 and 27). As the CPRD broadly represents the UK population, and SEER-Medicare includes a large, diverse population of older women diagnosed with breast cancer, results are generalisable to women diagnosed with ER/PR+ breast cancer in both the UK, US, and other developed populations due to the homogenous indication of endocrine therapy worldwide.

In the UK, ER/PR status was not available, but it is likely that breast cancers were ER/PR+ as such a diagnosis is a prerequisite of being prescribed endocrine therapies. The unavailability of ER/PR status also meant that we could not identify a group of untreated patients diagnosed with ER/PR+ breast cancer in the UK. Furthermore, CPRD captures prescriptions at the point of issue, but we do not know if prescriptions were filled, which could lead to potential misclassification of exposure. However, descriptive analyses using a 6 month grace period to define a continuous prescription indicated that 97% of women continued to be prescribed within 1 year of starting, 95% within 3 years, and 85% within 5 years.

In the US, the relatively high proportion of non-initiators of endocrine therapy (21%) in women with ER/PR+ was similar to the proportion of non-initiators reported in a previous study.[17] Reasons for non-initiation may include frailty, poor CVD preventative care, and high BMI, introducing possible residual confounding, especially given lack of information on smoking and BMI in US data. Although quantitative bias analyses suggest it is unlikely that residual confounding explains all of the large observed protective associations between tamoxifen use and several CVD outcomes, these associations may have been exaggerated, and the smaller observed associations between AI use and outcomes could be driven by such confounding. Comparisons to non-user groups are often more subject to confounding than comparisons to groups utilizing similar medications (i.e. AI vs tamoxifen).[37] Some caution should be taken in the interpretation of the comparisons with non-users, and these results warrant further investigation.

### Implications

Our results indicated no excess cardiotoxicities of endocrine therapy, other than the raised risk of VTE with tamoxifen. There was no evidence of raised risk of any CVD with AI use, including in stratified results restricted to those with prior CVD, suggesting that a history of cardiovascular disease should not be a contraindication for prescription of an AI. It is likely that AIs will continue to be the preferred endocrine therapy of choice in postmenopausal survivors of ER/PR+ breast cancer given its superior efficacy over tamoxifen. However, in women at a high CVD risk, it would be informative to know if any cardio-protective effects of tamoxifen might offset the superior reduction in risk of breast cancer recurrence with AIs. Further knowledge of the risk-benefit balance of the two endocrine therapies in terms of effects on cancer survival and cardiovascular outcomes in sub-groups of women would help guide individual-level prescribing decisions.

## CONCLUSION

Among postmenopausal women diagnosed with ER/PR+ breast cancer, we found convincing evidence of increased risks of several CVDs in AI compared with tamoxifen users. However, these associations appeared to be driven by protective effects of tamoxifen, rather than any toxic effects of AIs. There was no evidence that either drug type causally increased the risks of CVD outcomes, other than the known increased VTE risk with tamoxifen use. This information will help to inform and better understand the risk-benefit balance of these widely used endocrine therapies.

## Data Availability

Data from either the UK or US studies are not openly accessible, but can be gained through an application to the CRPD and SEER-Medicare respectively.

## FOOTNOTES

The corresponding author has the right to grant on behalf of all authors and does grant on behalf of all authors, an exclusive license (or non-exclusive for government employees) on a worldwide basis to the BMJ Publishing Group Ltd to permit this article (if accepted) to be published in BMJ editions and any other BMJPGL products and sub-licenses such use and exploit all subsidiary rights, as set out in our license.

## Contributions

Study designs were decided on by AM, JL, and KB. AM and SPH carried out data extraction. AM carried out all analyses. AM wrote the first draft. All authors contributed to further drafts and approved the final manuscript.

## Ethical approval

The UK study was approved by the ethics board at the London School of Hygiene and Tropical Medicine (project ID: 12025), and the US study was gained IRB approval at the University of North Carolina (study number: 17-1766)

## Transparency

The lead author affirms that the manuscript is an honest, accurate, and transparent account of the study being reported; that no important aspects of the study have been omitted; and that any discrepancies from the study as planned (and, if relevant, registered) have been explained.

